# Effect of improved spatial resolution on PET SUVR estimates in the LEADS study of sporadic early-onset Alzheimer’s disease

**DOI:** 10.1101/2025.01.13.25320481

**Authors:** Daniel R. Schonhaut, Renaud La Joie

## Abstract

**Background:** The Longitudinal Early-onset Alzheimer’s Disease Study (LEADS) recently transitioned from processing positron emission tomography (PET) scans at 8mm to 6mm resolution in early 2024. Concurrent with this improved spatial resolution, we revised our processing methods for defining reference regions used in amyloid and tau PET analyses, with the goal to maintain alignment with MRI-based PET processing in the Alzheimer’s Disease Neuroimaging Initiative (ADNI).

**Methods:** We describe the following changes to the LEADS MRI-based PET processing pipeline:

1. Transition from 8mm to 6mm PET scans.
2. Revised method to calculate a composite reference region scaling factor for longitudinal analysis of ^18^F-Florbetaben (FBB).
3. Revised method to define the eroded subcortical white matter region, which is used in longitudinal analyses of both FBB and ^18^F-Flortaucipir (FTP).
4. Corrected method to define the inferior cerebellar gray matter reference region used in cross-sectional analysis of FTP.

We evaluated these changes by comparing 8mm standardized uptake value ratios (SUVRs) processed with the original LEADS PET pipeline to 6mm SUVRs processed with the updated pipeline. This analysis was conducted in 564 FBB, 555 FTP, and 134 FDG scans acquired in LEADS participants at baseline between May 2018 and September 2023.

**Results:** We found near-perfect correlations across all tracers and reference regions (R^2^ > 0.99). However, 6mm SUVRs had greater dynamic range than 8mm SUVRs, with 8mm-to-6mm linear transformation slopes ranging from 1.08 to 1.22.

**Conclusions:** Our findings closely parallel ADNI’s experience transitioning to higher resolution PET. Amyloid PET Centiloid conversion equations, which were originally derived using 8mm data, should be re-estimated using 6mm data to maintain optimal harmonization. LEADS data users should not mix 6mm and 8mm PET data and should instead exclusively use 6mm PET data going forward, as these measures better reflect modern scanner capabilities while maintaining compatibility with ADNI. LEADS data users who are unsure of the origin or resolution of their PET values are encouraged to contact us for advice at.

## INTRODUCTION

PET processing in LEADS is designed to mimic the processing methods used by the Alzheimer’s Disease Neuroimaging Initiative (ADNI).^1^ An early step in ADNI’s processing entails smoothing PET scans to a uniform spatial resolution using Gaussian kernels that are separately estimated for each PET scanner at each clinical site. This smoothing step helps to harmonize PET scans reconstructed at different native resolutions, making the data more readily comparable across sites.

When LEADS launched in 2018, PET scans were smoothed to 8mm^3^ FWHM following the methods used by ADNI at the time. Then in 2022, ADNI began smoothing PET to 6mm instead of 8mm, a change that was made possible by improvement in the lowest resolution PET scanner used across ADNI clinical sites. The ADNI PET Core at UC Berkeley described this transition from 8mm to 6mm PET processing in a 2023 document uploaded to the Laboratory of Neuro Imaging (LONI) Image Data Archive (IDA),^2^ which serves as the central neuroimaging data repository for both LEADS and ADNI.

To maintain compatibility with ADNI, the LEADS PET Cores at UCSF and the University of Michigan recently transitioned away from processing PET scans at 8mm and adopted the same 6mm resolution as the new ADNI standard. All LEADS PET scans acquired prior to October 1, 2023 were reprocessed and reuploaded to LONI at 6mm. All LEADS PET scans acquired from October 1, 2023 onward are now processed and made available on LONI only at 6mm. Quantitative PET values in regions-of-interest (ROIs), which are uploaded to LONI by UCSF and shared with other LEADS investigators on a quarterly basis, are now derived exclusively from 6mm PET data. **It is still possible to download older, 8mm LEADS PET scans from LONI by conducting an advanced search for preprocessed PET scans with “Uniform Resolution” in the Image Description field. In contrast, all preprocessed PET scans at 6mm contain “Uniform 6mm Res” in the Image Description field**.

Concurrent with the change in PET resolution, we updated our MRI-based processing pipeline to correct several differences that we discovered between LEADS’ and ADNI’s methods for defining reference regions used to quantify amyloid and tau PET SUVRs. Here we describe these changes in detail and announce our public release of the complete, updated MRI-based PET processing pipeline used in LEADS, which can be downloaded from GitHub at https://github.com/rablabservice/leads_processing.

Using baseline LEADS PET data acquired between May 2018 and September 2023, we directly compared 8mm PET scans processed with our original pipeline to 6mm PET scans processed with our updated pipeline. We found that although 6mm and 8mm SUVR values were highly correlated (R^2^ > 0.99), SUVRs at 6mm had greater dynamic range than 8mm SUVRs, such that linear transformations from 8mm to 6mm had slopes significantly greater than 1. **It is therefore important not to mix PET values processed at 6mm and 8mm**. Our findings also imply that for amyloid PET, separate Centiloid conversion equations should be estimated for 6mm PET scans, as opposed to using the equations estimated on 8mm ADNI PET scans by Royse et al., 2021.^3^

## MATERIALS AND METHODS

**Scan inclusion**. We identified LEADS PET scans that were acquired between May 2018 and September 2023, the period during which both 6mm and 8mm pre-processed images were available for download from LONI. We limited our analysis to baseline PET scans for each tracer, and we excluded scans from participants who did not meet all LEADS inclusion criteria.^4^ We also excluded PET scans from subjects who were missing an acceptable MRI within one year of PET, along with PET scans that did not pass quality inspection checks. The final dataset consisted of 564 ^18^F-Florbetaben (FBB) scans, 555 ^18^F-Flortaucipir (FTP) scans, and 134 ^18^F-Fluorodeoxyglucose (FDG) scans (**Table 1**). Note that FDG was acquired only in control participants and patients assigned to the early-onset non-AD group (EOnonAD), as per LEADS protocol.

**Table 1.**
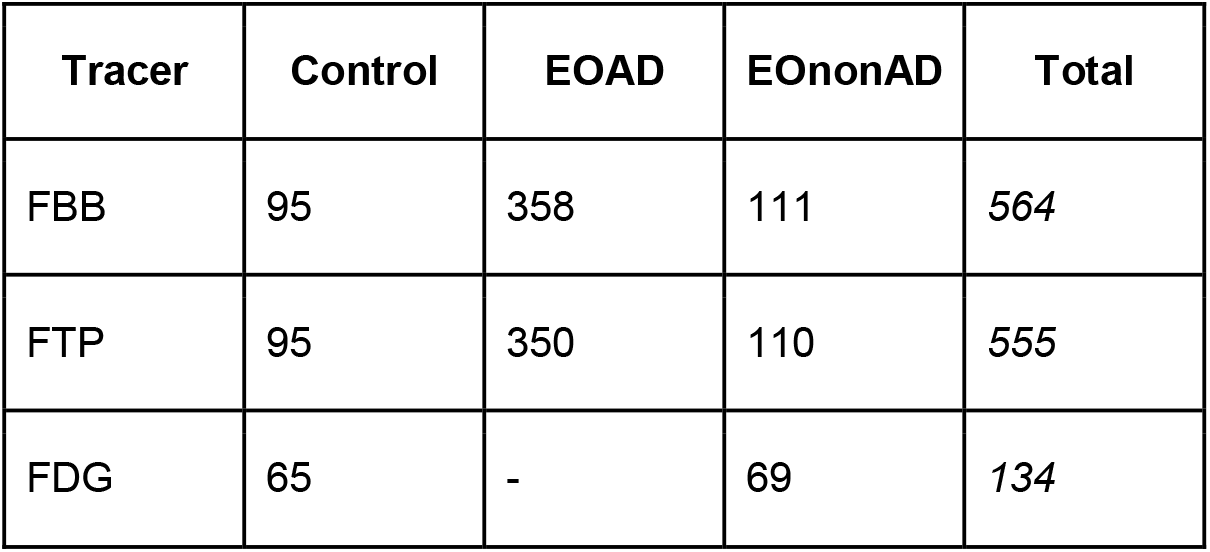
Number of LEADS participants included from each diagnostic group, for each PET tracer. The data included in this report is limited to one scan per patient; i.e. baseline data for each tracer. EOAD = Early-onset AD (visually FBB positive) patients. EAnonAD = Early-onset non-AD (visually FBB negative) patients.

### PET preprocessing

Reconstructed PET scans were uploaded to LONI by LEADS clinical sites and downloaded at the University of Michigan, where they were preprocessed using identical methods to those used in ADNI.^5^ Briefly, reconstructed PET frames were realigned and averaged over tracer-specific time windows (FBB: 90-110min, FTP: 75-105min, FDG: 30-60min), resliced to a uniform voxel size, and smoothed to a uniform resolution of 6mm^3^ or 8mm^3^ FWHM, respectively. These “Step 4” images were then uploaded back to LONI and labeled with the “Pre-processed” image type to distinguish these files from the PET images labeled “Original” that are uploaded by clinical sites.

### MRI-based PET processing

Preprocessed PET scans and T1-weighted MRIs were downloaded from LONI and processed at UCSF using one of two versions of our MRI-based processing pipeline. 8mm PET scans were processed using our original pipeline, while 6mm PET scans were processed using an updated version of our pipeline that was implemented in 2024 alongside our transition from 8mm to 6mm PET. Both pipeline versions follow the same sequence of core steps:

1. For each PET scan, we selected the closest available MRI that passed our quality inspection for suitable use in research.
2. MRIs were processed using FreeSurfer version 7.1.0, first by running the “recon-all” program and then by running “segmentBS.sh” to obtain a mask of the pons.
3. Preprocessed PET scans were rigid-body coregistered and resliced to their corresponding MRIs using Statistical Parametric Mapping version 12 (SPM12).
4. PET SUVRs were calculated in subject native MRI space using tracer-specific reference regions and target regions, as follows:
  a. Two SUVRs were calculated for each FBB scan: one using the FreeSurfer-defined whole cerebellum (aparc+aseg labels 7, 8, 46, and 47) as the reference region, and the second using a composite reference region spanning the whole cerebellum, brainstem (aparc+aseg label 16), and eroded subcortical white matter (see Methods section “Changes to the MRI-based PET processing pipeline” below). A single target region was defined across 40 left and right FreeSurfer ROIs; this is the “cortical summary” region used by ADNI to convert SUVRs to Centiloids (**Table S1**).^6^ Note that although our method for calculating the whole cerebellum and cortical summary regions did not change between the original and updated versions of our processing pipeline, our method for calculating the composite reference region did change, as described in the next section.
  b. Two SUVRs were calculated for each FTP scan: one using the inferior cerebellar gray matter as the reference region, and the other using an eroded subcortical white matter mask as the reference region. A single target region was defined, the “meta-temporal” ROI composed of FreeSurfer’s left and right amygdala, entorhinal cortex, parahippocampal gyrus, fusiform gyrus, and inferior and middle temporal cortices (aparc+aseg labels 18, 54, 1006, 1007, 1009, 1015, 1016, 2006, 2007, 2009, 2015, and 2016).^7^ While our method for calculating the meta-temporal region did not change between original and updated versions of our processing pipeline, our methods for calculating the inferior cerebellar gray matter and eroded subcortical white matter masks did change, as described in the next section.
  c. A single SUVR was calculated for each FDG scan, using the pons as the reference region and the same meta-temporal ROI that is used for FTP quantification as the target region.

### Changes to the MRI-based PET processing pipeline

The following changes to our original MRI-based PET processing pipeline were implemented in 2024 alongside our transition from 8mm to 6mm PET quantification. Each change was intended to bring PET processing methods in LEADS more in alignment with those used in ADNI, to correct a programming error in the original pipeline, or to correct an automated parcellation failure that we discovered in some highly atrophic MRIs, which had previously led to our excluding these data from analysis.

1. We revised our method to create the eroded subcortical white matter mask, which is used as a reference region for FTP and as part of the composite reference region for FBB.
  a. In our original pipeline, the eroded white matter mask was obtained by smoothing a binary mask of FreeSurfer’s cerebral white matter ROI (aparc+aseg labels 2 and 41) by 8mm^3^ FWHM, then binarizing voxels > 0.7 in the smoothed image.
  b. Our revised method works as follows:
    i. Create a subcortical white matter mask by combining aparc+aseg labels 2 and 41 and binarizing the output.
    ii. Zero-out voxels in the subcortical white matter mask with > 0.5 probability of being in the cerebrospinal fluid (CSF), as determined by the c3 tissue probability map from SPM12 segmentation. This cleaned version of the subcortical white matter mask is then used in place of the original subcortical white matter mask in subsequent steps. We added this step after noting that FreeSurfer occasionally mislabeled parts of the lateral ventricles as cerebral white matter on MRIs with severe ventricular enlargement. In most of these cases, SPM12 did not similarly mislabel CSF as white matter.
    iii. In SPM12, smooth the subcortical WM mask by 8mm^3^ FWHM.
    iv. Binarize voxels > 0.7 in the smoothed image to obtain the initial eroded subcortical white matter mask.
    v. Zero-out voxels in the initial eroded subcortical white matter mask that were not included in the subcortical white matter mask to obtain the final eroded subcortical white matter mask. This last step was not part of our original processing pipeline as it was not mentioned in the paper that defined the eroded subcortical white matter methodology.^5^ However, on consulting ADNI’s code, we found that this two-pass voxel removal procedure was implemented.
2. We revised our method to calculate a scaling factor for the composite reference region used in longitudinal FBB analyses. The scaling factor is the value calculated for each reference region, for each PET scan, that we divide all voxels in the Step 4, preprocessed PET image by to convert it to an SUVR image. In addition to changing how we create the eroded subcortical white matter mask (see #1 above), our original pipeline calculated a composite reference region scaling factor as the volume-weighted mean PET signal across the whole cerebellum, brainstem, and eroded subcortical white matter. However, after consulting with ADNI, we learned that their method for obtaining a scaling factor for the composite reference region calculates the mean PET signal within the whole cerebellum, brainstem, and eroded subcortical white matter separately, then takes an unweighted mean (i.e. not correcting for volume differences) across these three subregion values. Our revised method implements this same procedure.
3. To create the inferior cerebellar gray matter mask that is used as a reference region for FTP, the cerebellar SUIT atlas is nonlinearly transformed to native MRI space, where it is combined with a cerebellar gray matter mask defined from FreeSurfer-labeled aparc+aseg regions 8 and 47, as previously described.^8^ In our original processing pipeline, a version of the SUIT atlas in Montreal Neurological Institute (MNI) space was transformed to native MRI space while accidentally using trilinear interpolation to reslice voxels, whose final values were then recast from floating point to integer values. This unintended use of trilinear instead of nearest neighbor interpolation caused edge effects in which nonsensical atlas labels appeared along the borders between cerebellar subregions. In our updated pipeline, we use the SPM12 SUIT toolbox to create a subject-specific version of the SUIT atlas in native MRI space, avoiding edge effects from an improper interpolation algorithm. The rest of our method to define the inferior cerebellar gray matter mask is consistent with the procedure described in Baker et al., 2017.^8^

### Statistical analysis

For each PET tracer and reference region of interest, we used ordinary least-squares (OLS) univariate regression to calculate the slope and intercept that linearly transform 8mm to 6mm SUVRs, along with the 95% parametric confidence intervals for these estimates. For FBB and FTP scans, we also examined the linear relations between SUVRs calculated using cross-sectional versus longitudinal reference regions, separately for 6mm and 8mm PET scans. Results from these analyses are summarized in **Table 2** and **Figures 1-3**.

**Table 2.** Slope and intercept estimates are shown with 95% confidence intervals for the 8mm-to-6mm SUVR transformation equation for each PET tracer, reference region, and target region of interest. The last column shows the R^2^ for each univariate regression model fit.

| Tracer | Reference region | Target region | Slope | Intercept | $R^2$ |
| --- | --- | --- | --- | --- | --- |
| FBB | Whole cerebellum | Cortical Summary | 1.11 (1.11, 1.12) | -0.13 (-0.14, -0.13) | 0.9994 |
|  | Composite white matter | Cortical Summary | 1.09 (1.07, 1.10) | -0.08 (-0.09, -0.07) | 0.9919 |
| FTP | Inferior cerebellar gray matter | Meta-Temporal | 1.09 (1.09, 1.09) | -0.10 (-0.10, -0.09) | 0.9997 |
|  | Eroded subcortical white matter | Meta-Temporal | 1.22 (1.21, 1.23) | -0.21 (-0.23, -0.20) | 0.9956 |
| FDG | Pons | Meta-Temporal | 1.08 (1.06, 1.10) | -0.06 (-0.09, -0.04) | 0.9950 |

**Figure 1.**
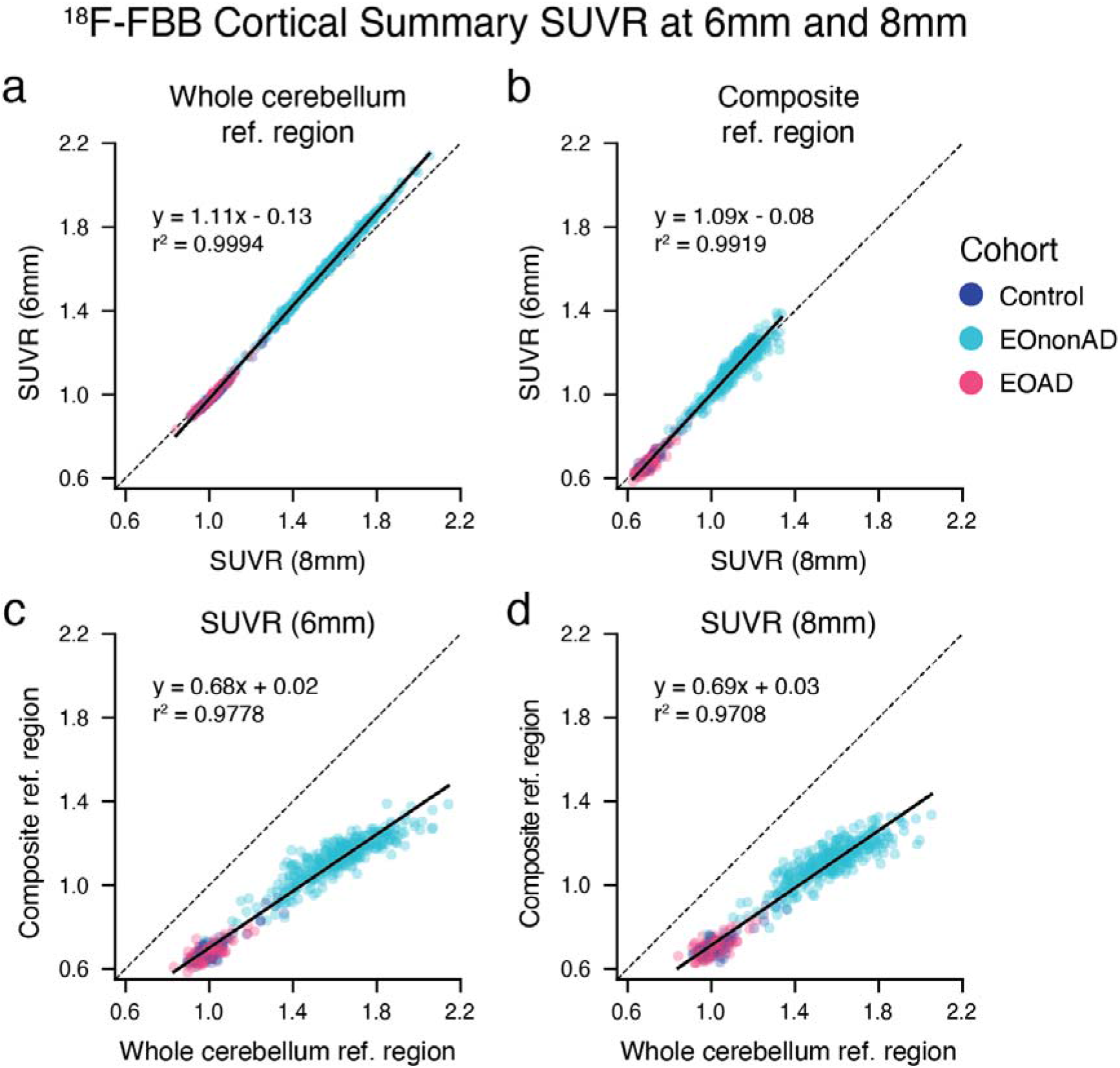
FBB SUVRs at 6mm versus 8mm resolution. (**a,b**) Scatterplots show the relationship between 6mm and 8mm SUVRs in the cortical summary target region scaled against the whole cerebellum (**a**) or composite reference region (**b**). (**c,d**) Scatterplots show the relationship between whole cerebellum and composite reference region-normalized SUVRs at 6mm (**c**) and 8mm (**d**). In all plots, solid lines show the OLS regression fit, and dashed lines show the identity line.

### Code availability

All code for the updated version of our MRI-based PET processing pipeline is freely available online at https://github.com/rablabservice/leads_processing. The GitHub repository includes a detailed README file with instructions on how to implement LEADS-like PET processing for those interested in adapting it.

## RESULTS

Across all PET tracers and reference regions, we found near perfect correlations between SUVRs calculated from 6mm versus 8mm PET scans (all R^2^ > 0.99). However, 6mm SUVRs consistently showed greater dynamic range than 8mm SUVRs, as evidenced by transformation slopes significantly greater than 1 for all tracer and reference region combinations (**Table 2**).

For FBB PET, 8mm SUVRs exceeded 6mm SUVRs at low values, while conversely, 6mm SUVRs exceeded 8mm SUVRs at high values (**Figure 1a,b**). 8mm-to-6mm regression lines crossed the identity line at an SUVR of 1.18 for the whole cerebellum reference region and 0.97 for the composite reference region. We also examined the relationship between whole cerebellum and composite reference region-estimated FBB SUVRs, separately at each PET resolution. 6mm and 8mm PET scans showed similarly high (R^2^ > 0.97) shared variances between the two reference regions and were described by similar transformation equations (**Figure 1c,d**).

For FTP PET, 8mm SUVRs were approximately equal to 6mm SUVRs at floor values, which were near an SUVR of 1 for each reference region (**Figure 2a,b**). However, 6mm SUVRs exhibited greater dynamic range than 8mm SUVRs. This was particularly true for SUVRs referenced against the eroded subcortical white matter, which increased by 1.22 at 6mm for every 1-step increase at 8mm. The slope of the 8mm-to-6mm regression for the inferior cerebellar gray matter reference region was notably lower, with 6mm SUVRs increasing by 1.09 for every 1-step increase at 8mm. When comparing SUVRs between the two reference regions at each resolution separately, we found that the shared variance between the reference regions was marginally greater at 6mm (R^2^ = 0.91) than at 8mm (R^2^ = 0.88). The slope of the linear transformation from inferior cerebellar gray matter to eroded white matter referenced SUVRs was greater at 6mm (0.46) than at 8mm (0.40) (**Figure 2c,d**).

**Figure 2.**
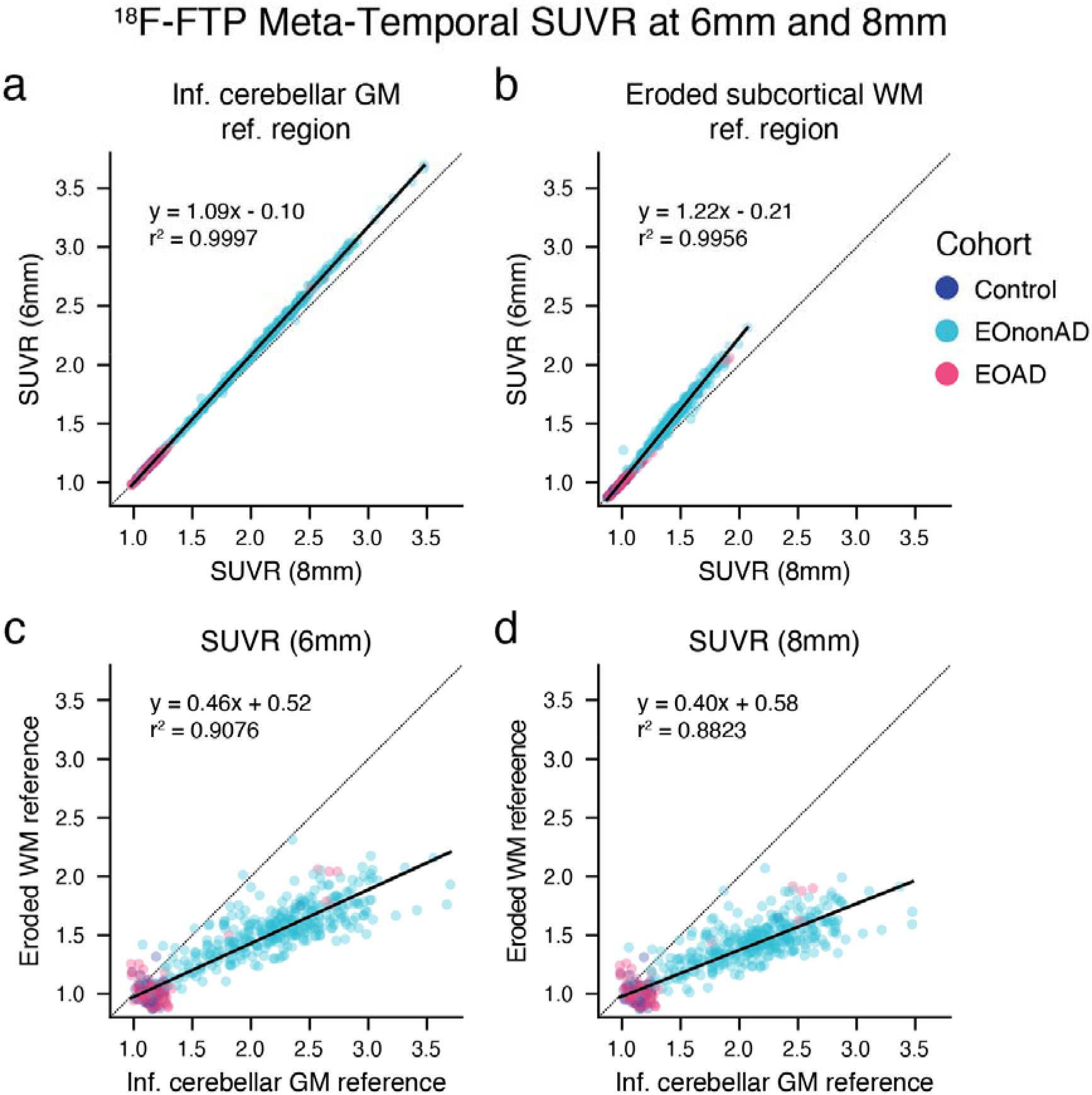
FTP SUVRs at 6mm versus 8mm resolution. (**a,b**) Scatterplots show the relationship between 6mm and 8mm SUVRs in the meta-temporal target region scaled against the inferior cerebellar gray matter (**a**) or eroded subcortical white matter reference region (**b**). (**c,d**) Scatterplots show the relationship between inferior cerebellar gray matter and eroded subcortical white matter reference region-normalized SUVRs at 6mm (**c**) and 8mm (**d**). In all plots, solid lines show the OLS regression fit, and dashed lines show the identity line.

For FDG PET, which used only the pons as a reference region, 6mm FDG SUVRs exceeded 8mm SUVRs across the full range of tracer values (**Figure 3**). The slope of the 8mm-to-6mm transformation equation (1.08) was comparable to that observed with the cross-sectional reference regions for FBB and FTP.

**Figure 3.**
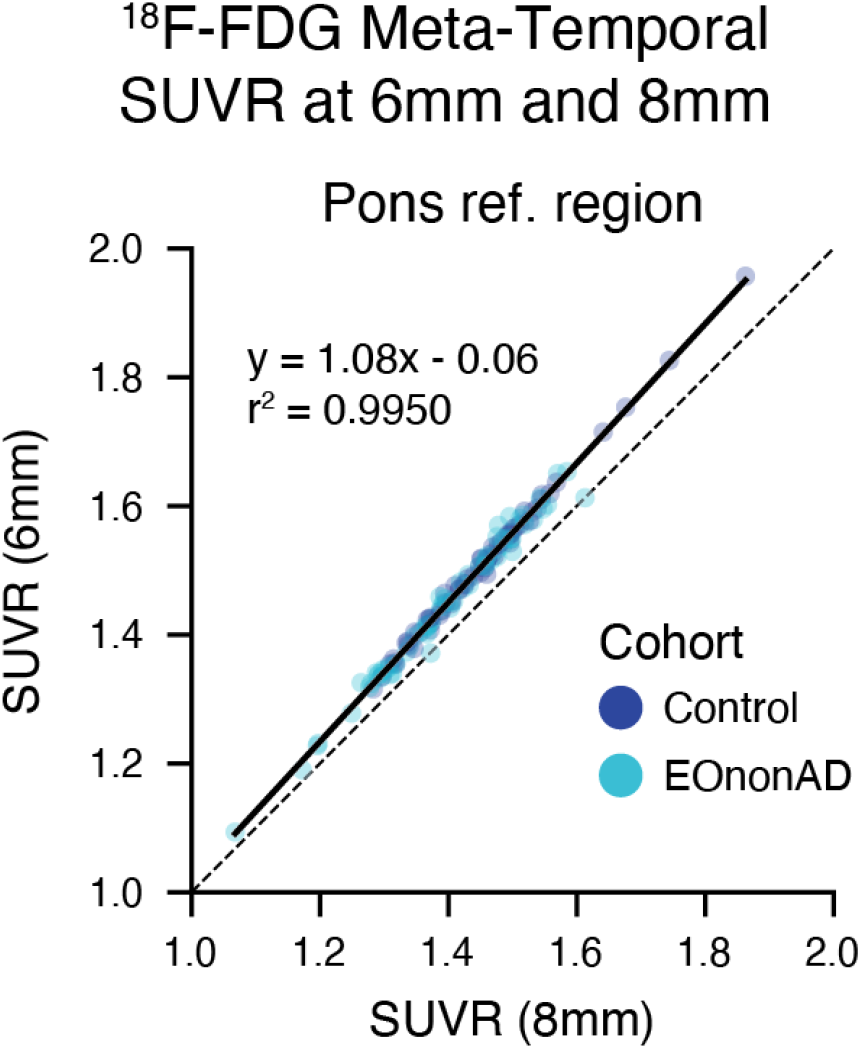
FDG SUVRs at 6mm versus 8mm resolution. Scatterplot shows the relationship between 6mm and 8mm SUVRs in the meta-temporal target region scaled against the pons. The solid line shows the OLS regression fit, and the dashed line shows the identity line.

## DISCUSSION

The transition from 8mm to 6mm PET processing in LEADS mirrors a similar change recently implemented in ADNI, reflecting improvements in PET scanner resolution across clinical sites. Alongside this transition, we updated our MRI-based PET processing pipeline to better align with ADNI’s methods, particularly in the creation of reference region masks used for SUVR calculation.

Our comparison of 6mm and 8mm PET data revealed highly consistent relationships across all tracers and reference regions. While 6mm and 8mm SUVRs were nearly perfectly correlated (R^2^ > 0.99), 6mm data consistently showed greater dynamic range, as evidenced by transformation slopes ranging from 1.08 to 1.22. The magnitude of this effect varied by tracer and reference region, with FTP SUVRs normalized to eroded subcortical white matter showing the largest difference between 6mm and 8mm resolutions. These findings closely parallel those reported by the UC Berkeley group in their analysis of ADNI PET data, where they similarly found transformation slopes greater than 1 and R^2^ values exceeding 0.96 for all PET tracer and reference region combinations they examined.^2^

Current Centiloid conversion equations used in LEADS and ADNI were derived from analyses of 8mm PET data by Royse et al., who established standardized equations to harmonize amyloid PET values for FBB and ^18^F-Florbetapir.^3^ However, our results indicate that 6mm and 8mm SUVRs, while highly correlated, are not directly comparable. Specifically, as 6mm SUVRs show greater dynamic range than 8mm SUVRs, applying the conversion equations estimated on 8mm PET data to 6mm data will yield less than optimally harmonized Centiloids, especially for highly positive scans where the greatest divergence between 6mm and 8mm SUVRs occurs. We recommend that new conversion equations be estimated using 6mm PET data for each amyloid PET tracer.

Given the systematic differences between 6mm and 8mm SUVRs, we also strongly recommend against combining PET measures at different resolutions in the same analysis. Instead, we advise LEADS data users to exclusively use 6mm PET data going forward, as these measures better reflect the improved resolution capabilities of modern PET scanners while maintaining compatibility with current ADNI methods.

## Data Availability

Data used for the analysis is available upon reasonable request from the authors.

https://leads-study.medicine.iu.edu/researchers/leads-data-request-application/

## CONTRIBUTIONS

DRS processed and analyzed the data and wrote the initial draft of the document. DRS and RLJ edited the document.

## DISCLAIMER

This document is presented by the authors as a service to LEADS data users and has not been evaluated by any external review process. Please direct correspondence to or to the LEADS PET Core at for general information.

## SUPPLEMENTARY MATERIALS

**Table S1.** The 40 FreeSurfer regions used to create the cortical summary target region mask for FBB SUVR quantification.

| FreeSurfer region | Left hemisphere label | Right hemisphere label |
| --- | --- | --- |
| caudalanteriorcingulate | 1002 | 2002 |
| caudalmiddlefrontal | 1003 | 2003 |
| inferiorparietal | 1008 | 2008 |
| inferiortemporal | 1009 | 2009 |
| isthmuscingulate | 1010 | 2010 |
| lateralorbitofrontal | 1012 | 2012 |
| medialorbitofrontal | 1014 | 2014 |
| middletemporal | 1015 | 2015 |
| parsopercularis | 1018 | 2018 |
| parsorbitalis | 1019 | 2019 |
| parstriangularis | 1020 | 2020 |
| posteriorcingulate | 1023 | 2023 |
| precuneus | 1025 | 2025 |
| rostralanteriorcingulate | 1026 | 2026 |
| rostralmiddlefrontal | 1027 | 2027 |
| superiorfrontal | 1028 | 2028 |
| superiorparietal | 1029 | 2029 |
| superiortemporal | 1030 | 2030 |
| supramarginal | 1031 | 2031 |
| frontalpole | 1032 | 2032 |

## REFERENCES

1. Landau, S. M. et al. Positron emission tomography harmonization in the Alzheimer’s disease neuroimaging initiative: A scalable and rigorous approach to multisite amyloid and tau quantification. Alzheimer’s Dement.: J. Alzheimer’s Assoc. (2024) doi:10.1002/alz.14378.

2. Lee, J. et al. PET 8mm-to-6mm Transformation Methods. (2023). ADNI Study Files document on LONI: https://ida.loni.usc.edu/explore/jsp/search/search.jsp?project=ADNI#studyFiles

3. Royse, S. K. et al. Validation of amyloid PET positivity thresholds in centiloids: a multisite PET study approach. Alzheimer’s Res. Ther. 13, 99 (2021).

4. Apostolova, L. G. et al. The Longitudinal Early-onset Alzheimer’s Disease Study (LEADS): Framework and methodology. Alzheimer’s Dement. 17, 2043–2055 (2021).

5. Landau, S. M. et al. Measurement of longitudinal β-amyloid change with 18F-florbetapir PET and standardized uptake value ratios. J. Nucl. Med.: Off. Publ. Soc. Nucl. Med. 56, 567–574 (2015).

6. Lee, J. et al. Amyloid PET Processing Methods. (2023). ADNI Study Files document on LONI: https://ida.loni.usc.edu/explore/jsp/search/search.jsp?project=ADNI#studyFiles

7. Jack, C. R. et al. Defining imaging biomarker cut points for brain aging and Alzheimer’s disease. Alzheimer’s Dement.: J. Alzheimer’s Assoc. 13, 205–216 (2017).

8. Baker, S. L., Maass, A. & Jagust, W. J. Considerations and code for partial volume correcting [18F]-AV-1451 tau PET data. Data Br. 15, 648–657 (2017).

